# Medical and Endovascular Treatment for Acute Large Vessel Occlusion with Mild Stroke: A Real-World Nationwide Registry

**DOI:** 10.1101/2025.02.05.25321767

**Authors:** Tingyu Yi, Shujuan Gan, Meihua Wu, Weifeng Huang, Dalong Sun, Yuehong He, Yanmin Wu, Dinglai Lin, Xiaohui Lin, Zhinan Pan, Yining Yang, Jinhua Ye, Caixia Li, Zhiting Chen, Mingzhu Huang, Xiufen Zheng, Xiaobin Li, Fenglong Lang, Qian Tan, Xiaojun Jin, Jianqiu Fu, Fuqiang Fan, Na Xu, Ya Shao, Yi Sui, Zhongrong Miao, Wenhuo Chen

## Abstract

**Background and Purpose:** EVT is a standard treatment for LVO in patients with an NIHSS score ≥6, but its role in mild stroke (NIHSS <6) remains uncertain. This study aims to explore the association between EVT and clinical outcomes in mild stroke.

**Methods:** This non-randomized cohort study prospectively enrolled consecutive mild stroke patients at 35 comprehensive stroke centers across 15 Chinese provinces from January 2020 to December 2023. Patients were categorized into pEVT or BMM groups, including those who received rEVT after deterioration. The primary outcome was an excellent outcome (mRS score of 0-1) at 90 days. Secondary outcomes included a good outcome (mRS score of 0-2) and mRS shift. Safety endpoints were mortality and sICH. Outcomes were compared between groups using multivariable logistic regression and IPTW.

**Results:** Finally, 307 patients were included. 110 received pEVT, and 190 received BMM. In the IPTW model, the pEVT group had higher rates of excellent (aOR=3.6, 95%CI=2.5-5.2) and good (aOR=4.0, 95%CI=2.5-5.6) outcomes, lower mortality (aOR=0.1, 95%CI=0.01-0.4), and a better 90-day mRS shift (aOR=0.2, 95%CI=0.1-0.3,), with similar sICH rates. Among the BMM group, 80 patients (42.1%) experienced early neurological deterioration (END), with 55 receiving rEVT. In the multivariable model, the pEVT group had higher rates of excellent (aOR=7.2, 95%CI=1.4-37.9) and good (aOR=4.1, 95%CI=1.2-14.8) outcomes and a better mRS shift (aOR=2.0, 95%CI=0.9-4.8).

**Conclusions:** Primary EVT significantly increases the likelihood of achieving an excellent outcome in mild stroke. Over 40% of mild stroke patients treated with BMM experienced END, and rEVT effectively improved the prognosis.

**Key Points:** *Question:* What is the optimal strategy for mild stroke with LVO?

*Findings:* This national prospective registry study demonstrated that pEVT significantly improved the prognosis of mild stroke patients with LVO compared to BMM. Approximately 40% of patients in the BMM group experienced END, and rEVT increased the likelihood of an excellent outcome in these patients.

*Meaning:* Primary EVT can significantly increase the probability of an excellent outcome in patients with mild stroke and LVO. Over 40% of patients with mild stroke treated with BMM experienced END, and rEVT effectively improved their prognosis.

## Introduction

Acute ischemic stroke (AIS) patients with a National Institutes of Health Stroke Scale (NIHSS) score <6 points is termed mild stroke, and mild stroke with large vessel occlusion (LVO) is not rare in the clinical scenario^1^. Nearly 40% of mild stroke with LVO have poor clinical outcomes when receiving conservative therapy^2^, and more proximal LVO is associated with early neurological deterioration (END) and poor clinical outcomes^3^. Endovascular therapy (EVT) can quickly open occluded arteries, restore blood flow to the ischemic brain, and is the standard treatment for AIS caused by LVO (AIS-LVO) with an NIHSS score ≥6 points^4^. However, whether mild stroke with LVO can benefit from EVT is controversial. Some studies favor EVT^2,5–9^ whereas other studies have shown that EVT is insignificant^10–17^, even causing harm by increasing the risk of symptomatic intracranial hemorrhage (sICH)^11,17^. Since the reperfusion state is associated with favorable clinical outcomes^18^, if the complications of EVT, such as dissection and hemorrhage, can be controlled, the benefit of EVT may be achieved in mild stroke with LVO.

In the modern EVT era, the device has become increasingly advanced, skills of neuro-interventionist have significantly improved, leading to higher successful reperfusion rate and fewer decreasing EVT-related complications. We hypothesize EVT can benefit patients with mild stroke and LVO. Therefore, we conducted this multicenter prospective registry study to evaluate the comparative efficacy and safety of primary EVT (pEVT) and best medical management (BMM) with or without rescue therapy EVT (rEVT) in treating mild stroke with LVO.

## Methods

### Design

The MILD MT(Endovascular Treatment for Mild Ischemic Stroke Due to Acute Large Vessel Occlusion in the Anterior Circulation) is a nationwide prospective registry involving consecutive patients aged 18 years or older with acute symptomatic, radiologically confirmed anterior circulation intracranial LVO. The study encompasses 35 comprehensive stroke centers across 15 provinces in China (Supplemental Figure I and Supplemental table I). To prevent selection bias, all participating centers were required to include all consecutive patients. Study centers had to perform at least 50 endovascular procedures annually, and all interventionists had to be certified in performing EVT for stroke patients with LVO. The study protocol was approved by the ethics committee of Zhangzhou Municipal Hospital, Fujian Medical University, China, and each subcenter. All patients or their legally authorized representatives provided signed, informed consent forms.

### Population

We included consecutive AIS patients who met the following criteria: (1) aged 18-80 years; (2) acute anterior circulation LVO confirmed by CTA or MRA; (3) onset-to -presentation time within 24 hours; (4) NIHSS score <6; and (5) ability to provide informed consent. For the pEVT group, EVT was initiated within 24 hours of estimated LVO onset. rEVT was permitted in the BMM group if END occurred. Exclusion criteria included: (1) premorbid modified Rankin Scale(mRS) score ≥1; (2) neuroimaging evidence of cerebral hemorrhage or edema; (3) lack of 90 - day follow - up data; (4) current pregnancy or lactation; (5) serious, advanced, or terminal illness; (6) incomplete baseline imaging or time data; (7) anticipated incomplete EVT; (8) multiple intracranial LVO without a clearly identifiable symptomatic vessel; (9) suspected or confirmed nonacute LVO; (10) poorly controlled hypertension (SBP >220 mmHg or DBP >120 mmHg); (11) baseline blood glucose <50 mg/dL or >400 mg/dL; (12) known bleeding tendencies; (13) seizures at onset or during the disease course affecting NIHSS assessment; or (14) aortic dissection.

### Data Collection

We collected demographic, clinical, and neuroimaging data from all eligible subjects, including age, sex, stroke risk factors (atrial fibrillation, hypertension, dyslipidemia, diabetes, and smoking), admission systolic blood pressure (SBP), admission NIHSS score, time from onset to imaging, and use of intravenous thrombolysis. Stroke etiology was defined according to the Trial of Org 10,172 in Acute Stroke Treatment (TOAST) classification^19^. The stroke occlusion location was categorized into the internal carotid artery (ICA), middle cerebral artery (MCA) (M1 or M2 segment), and tandem occlusion^20^. Brain tissue reperfusion was radiologically assessed immediately after the operation using the extended Thrombolysis in Cerebral Infarction (eTICI) scale. Successful reperfusion was defined as an eTICI score of ≥2b^18^.

### Outcome Measures

The primary outcome was an excellent outcome, which corresponds to a mRS score of 0–1 point. Secondary outcomes included a good outcome, represented by an mRS score of 0–2 points, and a shift in the mRS score at 90 days. Safety endpoints comprised mortality and symptomatic intracerebral hemorrhage (sICH).

Functional outcomes of the patients at 3 months were assessed using the mRS score, which ranges from 0 to 6 points, with higher scores indicating more severe disability. sICH was defined as any hemorrhage accompanied by an increase of ≥4 points in the total NIHSS score or an increase of ≥2 points in one NIHSS category, in accordance with the Heidelberg classification scheme^21^. Early neurological deterioration (END) was defined as a decrease of ≥4 points in the NIHSS score22 or a decrease of ≥2 points in a single motor item of the NIHSS within 7 days, which was not attributable to sICH.

### Statistical analysis

Continuous variables were expressed as mean ± SD or median (interquartile range) based on distribution normality, which was assessed by the Shapiro - Wilk test. These variables were compared using the Mann - Whitney U test or Student t - test. Categorical variables were reported as frequencies and percentages and compared using Pearson χ2 or Fisher exact test. To evaluate the association of pEVT with clinical outcomes, we conducted multivariable ordinal logistic regression analyses, and binary logistic regression was used for binary outcomes. Additionally, we performed multivariable ordinal logistic regression analyses to assess the association of rEVT with clinical outcomes in the BMM group with END. All analyses were adjusted for age, TOAST classification, hypertension, diabetes, atrial fibrillation, baseline NIHSS score, IV thrombolysis, and occlusion site. Statistical analysis was performed using SPSS Statistics 24.0 (IBM, Armonk, New York).

To estimate the average treatment effect of pEVT compared with BMM, we applied inverse probability of treatment weighting (IPTW). The process involved three steps: First, we generated propensity scores for pEVT using logistic regression models with age, TOAST classification, baseline NIHSS score, occlusion site, hypertension, diabetes, atrial fibrillation, and use of IV thrombolysis as predictors of treatment assignment. Second, we calculated weights (W) using the formula W = pEVT / PpEVT + (1 - pEVT) / (1 - PpEVT), where pEVT equals 1 for patients undergoing pEVT and 0 for those receiving BMM +/− rEVT, and PpEVT represents the estimated probability of receiving pEVT. Finally, these weights were incorporated into regression models to obtain marginal treatment effects. We assessed balance between the groups by measuring the absolute standardized difference, considering covariates with an absolute standardized difference of < 0.10 to be well - balanced.

### Subgroup Analysis

We tested treatment effect modification on the primary outcomes in different prespecified subgroups, including Age (≤70 vs. >70 years), Gender (Female vs. Male), Baseline NIHSS (≤3 vs. >3), Occlusion Site (ICA, M1, M2), and TOAST (CE, LAA, U). However, these analyses were considered exploratory only, as the study was not powered to draw conclusions from these data. The effect size of pEVT on the primary outcome measures in these subgroup analyses was assessed using weighted logistic regression, including the same weights obtained from the IPTW procedure. To account for multiple comparisons, we applied the Bonferroni correction to adjust the significance level for each individual hypothesis tested in the subgroup analyses. Otherwise, a significance level of P < 0.05 was set for the analyses, with all P values being 2 - sided. All statistical analyses were performed using R Software, v4.4.1.

## Results

### Patient selection and baseline characteristics

Among patients with AIS admitted to stroke centers between January 1, 2020, and December 2023, a total of 307 met the inclusion criteria. Of these patients, 117 were treated with pEVT, and 190 were treated with BMM. A total of 80 patients (42.1%) in the BMM group experienced early neurological deterioration (END). A total of 86.7% of the patients had large artery atherosclerosis (LAA), 13.0% had embolism, and 1.3% had an undetermined cause. A total of 99 patients in the pEVT group and 45 patients in the BMM group underwent CTP.

### Comparison of baseline characteristics and clinical outcomes between the pEVT and BMM +/− rEVT groups

The baseline characteristics of the patients are presented in Table 1. Compared with the BMM +/− rEVT group, patients in the pEVT group had a higher baseline NIHSS score (4 vs. 3.5 points), a lower NIHSS score at the severe stage (4 vs. 5 points), a higher prevalence of atrial fibrillation and/or heart valve disease (17.1% vs. 2.6%), a higher incidence of tandem occlusion (14.5% vs. 6.8%), a lower proportion of large artery atherosclerosis (LAA) (70.9% vs. 94.7%, p < 0.001), a higher proportion of embolism (25.6% vs. 5.3%, p < 0.001), and greater hypo - perfused and mismatch volumes (102 ml vs. 70 ml; 93 ml vs. 59 ml, p = 0.007).

**Table 1.**
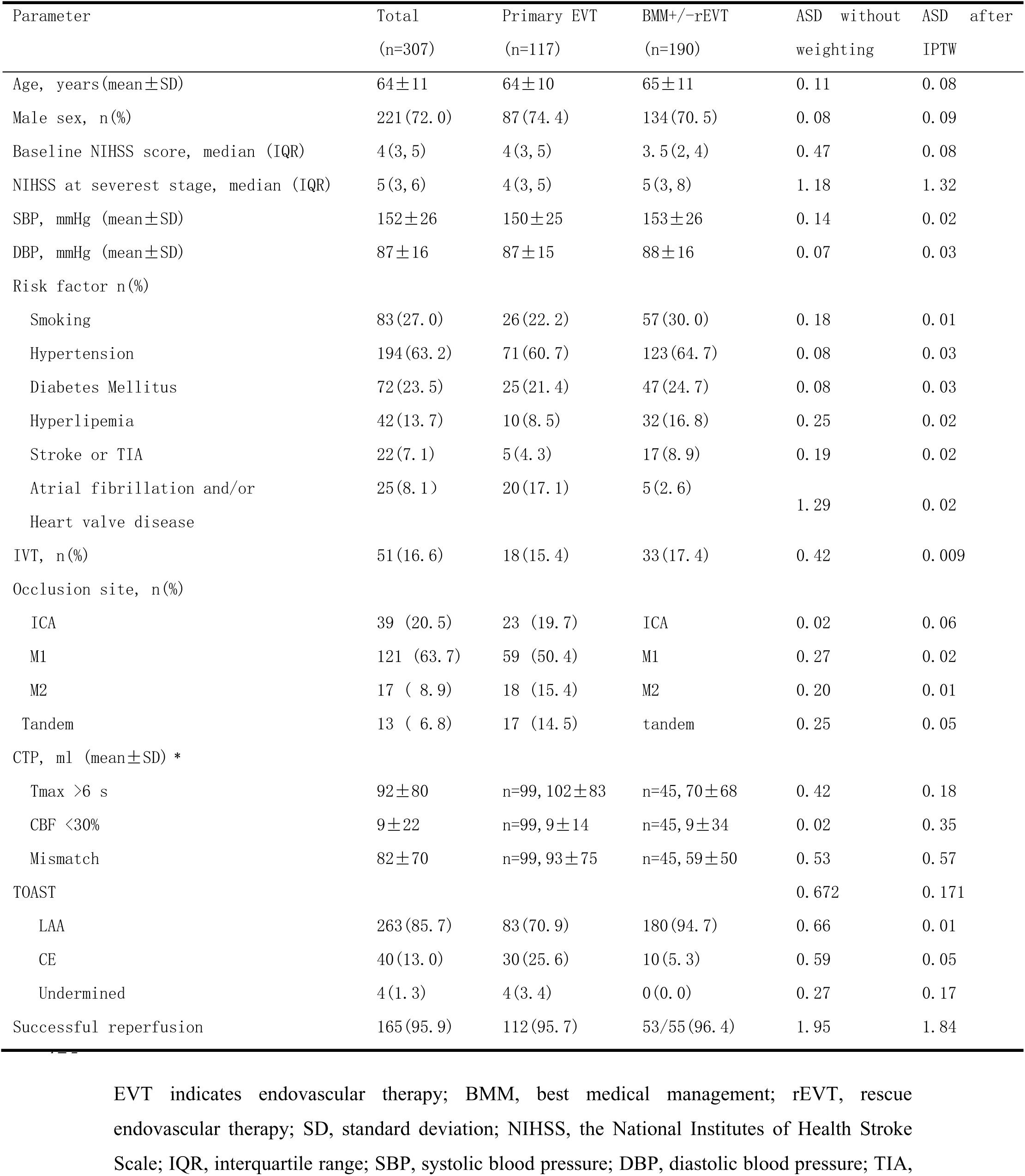
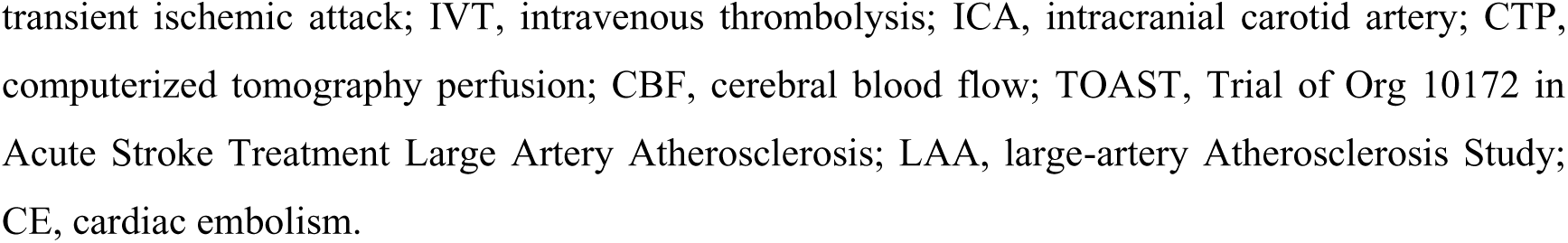
Demographics and characteristics of the included patients stratified by treatment allocation.

| Parameter | Total<br>(n=307) | Primary EVT<br>(n=117) | BMM+/-rEVT<br>(n=190) | ASD without<br>weighting | ASD after<br>IPTW |
| --- | --- | --- | --- | --- | --- |
| Age, years (mean±SD) | 64±11 | 64±10 | 65±11 | 0.11 | 0.08 |
| Male sex, n(%) | 221 (72.0) | 87 (74.4) | 134 (70.5) | 0.08 | 0.09 |
| Baseline NIHSS score, median (IQR) | 4 (3, 5) | 4 (3, 5) | 3.5 (2, 4) | 0.47 | 0.08 |
| NIHSS at severest stage, median (IQR) | 5 (3, 6) | 4 (3, 5) | 5 (3, 8) | 1.18 | 1.32 |
| SBP, mmHg (mean±SD) | 152±26 | 150±25 | 153±26 | 0.14 | 0.02 |
| DBP, mmHg (mean±SD) | 87±16 | 87±15 | 88±16 | 0.07 | 0.03 |
| Risk factor n(%) |  |  |  |  |  |
| Smoking | 83 (27.0) | 26 (22.2) | 57 (30.0) | 0.18 | 0.01 |
| Hypertension | 194 (63.2) | 71 (60.7) | 123 (64.7) | 0.08 | 0.03 |
| Diabetes Mellitus | 72 (23.5) | 25 (21.4) | 47 (24.7) | 0.08 | 0.03 |
| Hyperlipemia | 42 (13.7) | 10 (8.5) | 32 (16.8) | 0.25 | 0.02 |
| Stroke or TIA | 22 (7.1) | 5 (4.3) | 17 (8.9) | 0.19 | 0.02 |
| Atrial fibrillation and/or<br>Heart valve disease | 25 (8.1) | 20 (17.1) | 5 (2.6) | 1.29 | 0.02 |
| IVT, n(%) | 51 (16.6) | 18 (15.4) | 33 (17.4) | 0.42 | 0.009 |
| Occlusion site, n(%) |  |  |  |  |  |
| ICA | 39 (20.5) | 23 (19.7) | ICA | 0.02 | 0.06 |
| M1 | 121 (63.7) | 59 (50.4) | M1 | 0.27 | 0.02 |
| M2 | 17 (8.9) | 18 (15.4) | M2 | 0.20 | 0.01 |
| Tandem | 13 (6.8) | 17 (14.5) | tandem | 0.25 | 0.05 |
| CTP, ml (mean±SD) * |  |  |  |  |  |
| Tmax >6 s | 92±80 | n=99, 102±83 | n=45, 70±68 | 0.42 | 0.18 |
| CBF <30% | 9±22 | n=99, 9±14 | n=45, 9±34 | 0.02 | 0.35 |
| Mismatch | 82±70 | n=99, 93±75 | n=45, 59±50 | 0.53 | 0.57 |
| TOAST |  |  |  | 0.672 | 0.171 |
| LAA | 263 (85.7) | 83 (70.9) | 180 (94.7) | 0.66 | 0.01 |
| CE | 40 (13.0) | 30 (25.6) | 10 (5.3) | 0.59 | 0.05 |
| Undermined | 4 (1.3) | 4 (3.4) | 0 (0.0) | 0.27 | 0.17 |
| Successful reperfusion | 165 (95.9) | 112 (95.7) | 53/55 (96.4) | 1.95 | 1.84 |

EVT indicates endovascular therapy; BMM, best medical management; rEVT, rescue endovascular therapy; SD, standard deviation; NIHSS, the National Institutes of Health Stroke Scale; IQR, interquartile range; SBP, systolic blood pressure; DBP, diastolic blood pressure; TIA,
transient ischemic attack; IVT, intravenous thrombolysis; ICA, intracranial carotid artery; CTP, computerized tomography perfusion; CBF, cerebral blood flow; TOAST, Trial of Org 10172 in Acute Stroke Treatment Large Artery Atherosclerosis; LAA, large-artery Atherosclerosis Study; CE, cardiac embolism.

The comparison of clinical outcome is shown in Table 2. Multivariable analysis revealed that patients in the pEVT group had higher rates of excellent and good clinical outcomes (77.8% vs. 56.8%, aOR = 3.4, 95%CI = 1.8 - 6.3, p < 0.001; 86.3% vs. 71.1%, aOR = 2.3, 95%CI = 1.7 - 7.3, p < 0.001), lower mortality (0.9% vs. 3.7%, aOR = 0.1, 95%CI = 0.01 - 1.1, p < 0.037), and a better mRS shift (0 vs. 1, aOR = 0.2, 95%CI = 0.1 - 0.4, p < 0.001) compared with those in the BMM +/− rEVT group.

**Table 2.** Clinical outcomes of the included patients in the primary EVT and BMM+/− rEVT groups.

| Outcome | pEVT | BMM+/-rEVT | Unadjusted model |  | Multivariable model |  | IPTW model |  |
| --- | --- | --- | --- | --- | --- | --- | --- | --- |
|  | n=117 | n=190 | OR (95%CI) | P | aOR (95%CI) | P | aOR (95%CI) | P |
| mRS 0-1 at 90d | 91 (77.8) | 108 (56.8) | 2.6 (1.5-4.4) | <0.001 | 3.4 (1.8-6.3) | <0.001 | 3.6 (2.5-5.2) | <0.001 |
| mRS 0-2 at 90d | 101 (86.3) | 135 (71.1) | 2.3 (1.3-4.3) | 0.008 | 3.5 (1.7-7.3) | <0.001 | 4.0 (2.5-6.6) | <0.001 |
| mRS shift at 90d | 0 (0, 1) | 1 (0, 3) | 0.3 (0.2-0.5) | <0.001 | 0.2 (0.1-0.4) | <0.001 | 0.2 (0.1-0.3) | <0.001 |
| Safety outcome |  |  |  |  |  |  |  |  |
| sICH | 2 (1.8) | 6 (4.0) | 1.4 (0.4-4.6) | 0.611 | 1.0 (0.2-4.1) | 0.467 | 1.3 (0.5-2.9) | 0.570 |
| Mortality | 1 (0.9) | 7 (3.7) | 0.2 (0.02-1.9) | 0.166 | 0.1 (0.01-1.1) | 0.037 | 0.1 (0.01-0.4) | 0.002 |

Multivariable models adjust for age, baseline NIHSS, hypertension, diabetes, arterial fibrillation, IVT, TOAST, occlusion site; pEVT, primary endovascular therapy; BMM, best medical management; rEVT, rescue endovascular therapy; END, early neurological deterioration; mRS: modified Rankin scale; sICH, symptomatic intracranial hemorrhage.

### IPTW Model

The baseline demographics and clinical characteristics were well-balanced. Notably, computed tomography perfusion (CTP) parameters were not included in the IPTW sample creation, as only 24% of patients in the BMM group and 85% of patients in the pEVT group underwent CTP (Table 2). Consistent with the multivariable model - adjusted analysis, the IPTW model revealed that patients in the pEVT group had higher rates of excellent and good clinical outcomes (aOR = 3.6, 95%CI = 2.5 - 5.2, p < 0.001; aOR = 4.0, 95%CI = 2.5 - 5.6, p < 0.001), lower mortality (aOR = 0.1, 95%CI = 0.01 - 0.4, p = 0.002), and a better mRS shift (aOR = 0.2, 95%CI = 0.1 - 0.3, p < 0.001) compared with those in the BMM +/− rEVT groups. The distribution of the 90 - day mRS score by treatment allocation is illustrated in Figure 1.

**Figure 1.**
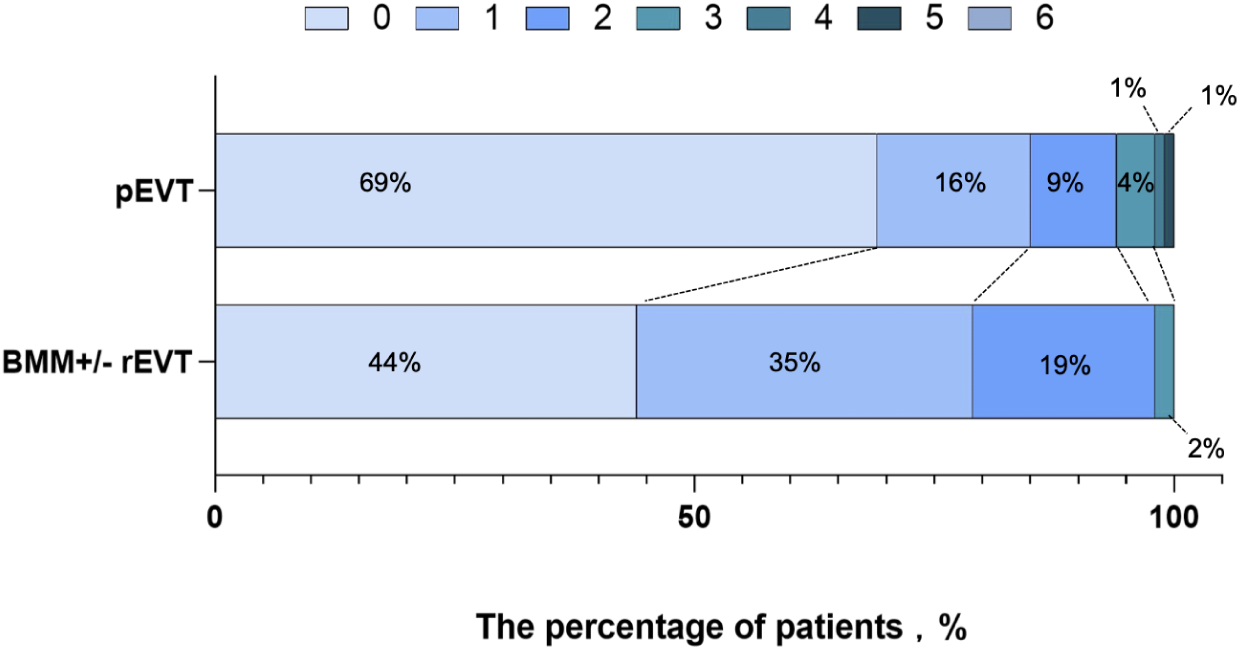
Distribution of the mRS score at 90 days stratified by treatment allocation mRS indicates modified Rankin Scale; pEVT primary endovascular therapy; BMM, best medical management.

### Comparison of the clinical outcomes of patients with END between the BMM groups with or without rEVT

n the BMM group, END occurred in 80 patients, 55 of whom received rEVT. The clinical outcome comparison between the two groups is shown in Supplemental Table II. Compared with END patients in the BMM without rEVT group, those in the rEVT group had higher rates of excellent and good outcomes (36.4% vs. 8.0%, aOR = 7.2, 95%CI = 1.4 - 37.9, p = 0.020; 47.3% vs. 20.0%, aOR = 4.1, 95%CI = 1.2 - 14.8, p = 0.030) and a better mRS shift (aOR = 2.0, 95%CI = 0.9 - 4.8, p = 0.010). The distribution of the 90 - day mRS score by treatment allocation is illustrated in Figure 2.

**Figure 2.**
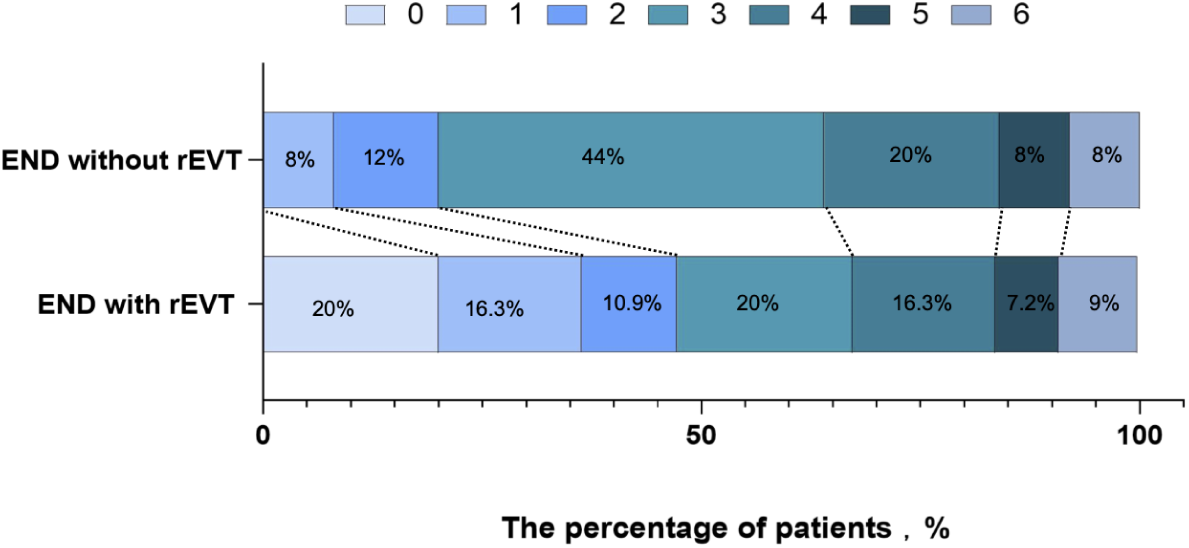
Distribution of the mRS score at 90 days patients with END stratified by treatment allocation mRS indicates modified Rankin Scale; END, early neurological deterioration; rEVT, rescue endovascular therapy.

### Comparison of the clinical outcomes of patients without END between the BMM and pEVT groups

In the BMM group, 110 patients without END were included. Compared with the BMM without END group, fewer patients in the pEVT group had a good prognosis (86.3% vs. 98.2%, aOR = 0.2, 95%CI = 0.34 - 0.70, p = 0.015), but there were similar excellent prognosis rates (79.1% vs. 77.8%, p = 0.808), sICH occurrence (1.8% vs. 3.4%, p = 0.506), and mortality (0% vs. 0.9%, p = 0.988). The distribution of the 90 - day mRS score by treatment allocation is illustrated in Supplemental Figure II.

### Subgroup Analyses

The relatively small sample size limited the power of the subgroup analyses. All subgroup analyses except for the age >70 years, baseline NIHSS ≤3, and MCA occlusion subgroups favored pEVT (Figure 3).

**Figure 3.**
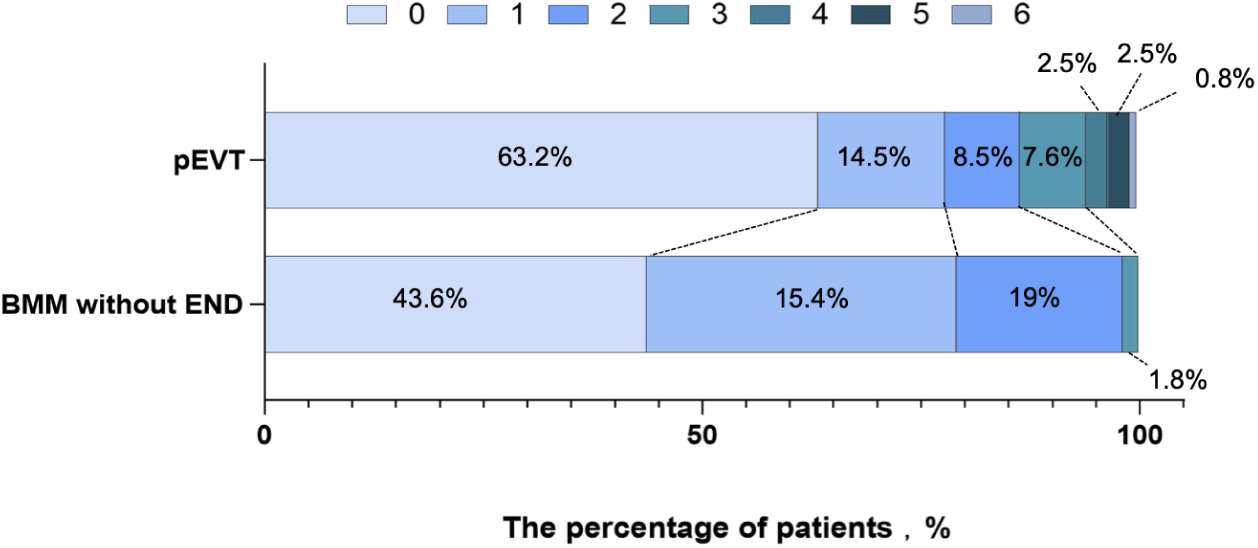
Treatment effect size of pEVT on excellent clinical outcome across different subgroups. pETV, indicates primary endovascular therapy; aOR, adjusted odd ratio; NIHSS, The National Institutes of Health Stroke Scale; TOAST, Trial of Org 10172 in Acute Stroke Treatment Large Artery Atherosclerosis; LAA, large-artery Atherosclerosis Study; CE, cardiac embolism, ICA, intracranial carotid artery, BMM, best medical management.

## Discussion

The main findings of our study can be summarized as follows: First, pEVT could improve the prognosis of mild stroke patients with LVO compared with the BMM group. Second, nearly 40% of patients who received BMM experienced END, and rEVT increased the probability of achieving an excellent outcome in patients with END. Third, pEVT did not improve the prognosis of mild stroke patients compared with BMM patients without END.

The benefit of EVT for mild stroke patients with LVO remains uncertain. A review article encompassing 10 studies that compared EVT with BMM in this patient population^23^, found that approximately 6 studies reported no benefit of VET for mild stroket^10–14,17^. Moreover, the meta-analysis^11^, which included 11 observational studies comprising 2019 patients with mild stroke treated with EVT and 3171 patients treated with BMM, revealed the similar results. The possible reasons for the negative results of these studies are as follows: small sample sizes, with the number of patients receiving EVT ranging from 10 to 54^12,13,17^; included of AIS patients caused by posterior circulation LVO^17^; a significant number of patients with medium distal vessel occlusion were included in the BMM group^11^; and successful reperfusion rates below 85%^11–14,17^. The reasons why our study yielded positive results may include the following: a. relatively large sample size; b. inclusion of only patients with anterior circulation LVO; c. exclusion of patients with medium distal vessel occlusion; d. a high successful reperfusion rate; e. a large mismatch volume on computerized tomography perfusion (CTP) in our cohort, which makes patients more likely to benefit from EVT, as reported in a previous study^24^; f. a high proportion of patients with intracranial artery atherosclerosis (ICAS), which is associated with good collateral circulation; and. g inclusion of patients admitted to comprehensive stroke centers, where EVT skills are proficient, and the risk of procedure - related complications can be minimized.

The occurrence of END caused by ischemic factors in mild stroke patients with LVO is relatively common, with incidence rates ranging from 20%^25^ to nearly 50%^26,27^. The causes of END include ischemia, hemorrhage and edema^28^, and ischemic END is twice as common as sICH^29^. The pathological mechanisms of ischemic END encompass artery-to-artery embolism, ischemic progression, artery re-occlusion, thrombus extension, and LAA or ICAS, all of which are independent predictors of END^28^. In addition to etiology, the occlusion site and thrombus length are also related to the occurrence of END^3^. Our study revealed that approximately 40% of patients experienced END, which was relatively high rate. This may be attributed to the the broad definition of END used in our study, where an increase of ≥2 points in a single motor item on the NIHSS was also considered indicative of END; Additionally, the observation period in our study was lengthy (7 days), and the risk of END increases with longer observation periods^25^. ICAS was the primary cause in our study and is an independent predictor of END^28^. Moreover, over 90% of patients in our study had ICA or MCA occlusion, which are also independent predictors of END^3^.

Once END occurred, the prognosis for patients managed medically was poor, with only 20% achieving functional independence, similar to previous findings^26^. However, this differed from Pierre Seners’ study, potentially due to the different proportions of M2 occlusion patients included (12% in our study vs. over 50% in theirs)^3^. Previous studies have demonstrated that rEVT is associated with better clinical outcomes in mild stroke patients with END^3,25^ which aligns with our findings. Our study also found that nearly 80% of patients with mild stroke without END managed medically had excellent outcomes. A higher rate of good outcomes was observed in patients without END receiving BMM compared to those receiving pEVT. These findings suggest that patients with mild stroke at risk of END may benefit most from EVT. Therefore, clinicians should focus on identifying predictors of END in mild stroke patients to select the most appropriate candidates for EVT. Previous research has shown that the mismatch volume detected by CTP is related to the occurrence of END, indicating that mild stroke patients with LVO selected by brain perfusion may benefit from EVT, a hypothesis that needs to be confirmed by the ongoing MILD MT tria (NCT06179017).

### Limitations

This study has several limitations. First, as a non-randomized study, it shares all the inherent limitations of such designs. The reasons for clinicians to choose a specific treatment option are more complex and cannot be captured by a prospective observational study. IPTW or multivariable analyses can adjust systematic differences between treatment groups to minimize the influence. Second, only stroke centers capable of providing EVT were included in our registry, which may have led to an overestimation of END occurrence. Third, only a small proportion of mild stroke patients underwent CTP, limiting our ability to fully explore the relationship between CTP parameters and the prognosis of mild stroke. Fourth, due to the registry nature of the study, no unified treatment protocol was specified, and the different practices of treating physicians at various centers might have influenced treatment decisions. Fifth, data on the time from presentation to END for patients receiving rEVT were not available for analysis. Sixth, the generalizability of the findings may be limited since the study was conducted only in China, where ICAS is prevalent.

## Conclusion

Primary EVT can improve the prognosis of mild stroke patients with LVO. Almost 40% of mild stroke patients experience END, and rEVT can enhance clinical outcomes in this patient group. These findings require confirmation by future randomized controlled trials (RCTs).

## Data Availability

Anonymized data will be shared upon request to any qualified investigator.

## Disclosures

All the authors declare no potential conflicts of interest.

## Data Sharing Statement

No additional unpublished data are available.

## Sources of Funding

Dr. Wenhuo Chen received grants from the Natural Science Foundation of Fujian Province (Grant No. 2022J01123162) and Dr. Tingyu Yi received grants from Beijing Health Promotion Association (Grant No. BHPA2021IN002) and National Health Commission Capacity Building and Continuing Education Center (Grant No. GWJJ20221003).

## Notes

### Competing Interest Statement

The authors have declared no competing interest.

### Author Declarations

The study protocol was approved by the ethics committee of Zhangzhou Municipal Hospital, Fujian Medical University, China, and each subcenter. All patients or their legally authorized representatives provided signed, informed consent forms.

